# Retrospective cohort analysis of surgical management for congenital diaphragmatic eventration in children

**DOI:** 10.1101/2024.02.15.24302855

**Authors:** Khalid Alzahrani, Lymeymey Heng, Naziha Khen-Dunlop, Nicoleta Panait, Erik Hervieux, Lucie Grynberg, Olivier Abbo, Frédéric Hameury, Frédéric Lavrand, Olivier Maillet, Aurore Haffreingue, Anne Lehn, Stephan De Napoli Cocci, Edouard Habonimana, Jean-Luc Michel, Louise Montalva, Quentin Ballouhey, Arnaud Fotso Kamdem, Jean-François Lecompte, Antoine Liné, Anna Poupalou, Pierre Meignan, Loren Deslandes, Guillaume Podevin, Françoise Schmitt

## Abstract

**Aim:** To compare the different surgical approaches for treating congenital diaphragmatic eventration (CDE) in children.

**Methods:** Retrospective data analysis of a multicentric cohort of pediatric patients operated on for CDE between 2010 and 2021, with comparison of the different surgical approaches and their outcomes.

**Results:** 112 patients, aged 12 [5–21] months, were operated on for CDE. Diaphragmatic plication was performed using thoracoscopy or RATS in 69 (62%) cases, postero-lateral thoracotomy (PLT) in 15 (13%), and using an abdominal approach in 28 (25%). Relief of symptoms was obtained in 88% of the cases and improvement in the diaphragmatic level in 90%. We recorded 31 postoperative complications (28%) and eight recurrences (7%). Compared to other approaches, PLT multiplied the duration of intravenous analgesia by three (96 vs 36h, p<0.0001) and the length of hospital stay by two (8 vs 4d, p=0.002). RATS provided more perioperative hepatic injuries and equivalent short-term results than thoracoscopy, but all five patients remained symptomatic and two of them experienced chest wall deformities.

**Conclusion:** Diaphragmatic plication via a minimally invasive thoracic approach may be the best treatment option for cases of symptomatic CDE. Further is required to confirm that RATS is, at least, not inferior to thoracoscopy.

ClinicalTrial NCT04862494, 2021-04-28;

## Introduction

Congenital diaphragmatic eventration (CDE) is a rare diaphragmatic malformation affecting about five out of every 10,000 live births [1], leading to an ascent of the diaphragmatic dome in the chest and to a paradoxical motion of the diaphragm. Most patients present with pulmonary and/or digestive symptoms and are often treated by a surgical diaphragmatic plication [2–4]. Until recently, a left transverse laparotomy had been recommended for left-sided CDE and a postero-lateral thoracotomy for right-sided procedures [5], both with good results in the short and long terms [1]. But the development of mini-invasive surgical approaches (e.g., video-assisted surgery, robotic surgery) has improved surgical recovery [6] without negatively impacting long-term outcomes, which are usually described as excellent [1,6,7].

In a previous study [8], we have recently described the clinical presentation of our whole cohort of patients and analyzed the reasons leading to surgical or conservative management of CDE. In the present study, we now aimed to determine which surgical treatment of CDE should be preferred by exploring the immediate and long-term outcomes and complications. Our secondary objectives were to identify potential predictors for surgical approaches’ choice, to investigate specific surgical risk factors for eventration recurrence, and to present our first cases of robotic plication.

## Materials and Methods

### 1) Subjects

Data were extracted from a retrospective multicenter cohort study on CDE management conducted in 22 pediatric surgery departments between January 1, 2010, and August 31, 2021 [8]. We included all patients under the age of 16 who had undergone operations for CDE. Patients with diaphragmatic hernia and diaphragmatic eventration secondary to surgery, obstetrical trauma, or oncological causes were excluded from analysis, as were all patients whose representatives were opposed to their participation in this study. Our research was conducted in accordance with the principles of the Declaration of Helsinki, obtained the agreement of our local ethics committee (n°2021-013) and of the French National Commission on Informatics and Liberty CNIL (Commission Nationale de l’Informatique et des Libertés, authorization ar21-001v0), and has been registered in ClinicalTrials.gov (NCT04862494).

Data recorded from medical charts included preoperative demographic and clinical symptoms at diagnosis and results from medical imaging studies on thoracic radiographs, CT scans, and MRIs when available. Perioperative data included the surgical approach, type of repair, material used, and the occurrence of complications, as well as early postoperative events. Clinical outcomes were assessed on postoperative recovery or the persistence of symptoms, on the radiographic level of the diaphragmatic dome, and on the occurrence of complications, the need for re-intervention or hospital stays. Indications for surgery included all clinical or radiological findings leading surgeons to propose CDE plication as a mean to improve the patient’s condition. As well, all postoperative events considered as abnormal during the postoperative course and associated to surgery were registered as complications.

### 2) Outcome measures

The primary outcome was to compare the different surgical approaches, classified as abdominal repair (open or laparoscopic), postero-lateral thoracotomy (PLT), and thoracic mini-invasive thoracoscopic surgery (MITS, including thoracoscopy and robot-assisted thoracic surgery). Secondary outcomes were to provide the initial results of a few cases of robotic surgery for CDE, to look for potential predictive factors for surgical approaches’ choice, and to identify surgical risks of recurrence.

### 3) Statistical methodology

Statistical analysis was performed using GraphPad Prism 8.0.2 and IBM-SPSS 29.0.0.0 for Windows. All tests were 2-sided and the statistical level for significance was set at p < 0.05. Patients’ characteristics were described as median with interquartiles for continuous variables, and as percentages for qualitative variables. Two-by-two comparative analysis of quantitative variables was performed using the non-parametric Mann-Whitney test after failing to pass a Shapiro-Wilk normality test and of qualitative variables with the Fisher’s exact test. Comparisons between multiple groups were made using the Kruskall-Wallis test for quantitative data and a Pearson’s Chi^2^ test for qualitative variables. Association analysis was performed on potentially relevant variables (p-value under 0.20 on bivariate analysis) using logistic regression, with a backward stepwise likelihood-ratio test, on the software. Data were then expressed as odds ratios (OR) with 95% confidence intervals.

## Results

### 1) Description of the patients

During the 12-year recruitment period, 112 patients underwent surgical treatment for CDE in the 22 pediatric surgery departments that participated in this study. The median age at diagnosis was eight [1–16] months with extremes going from antenatal diagnosis to 15.8 years, and 34 patients were diagnosed after 12 months (30.4%). Seventy-six (67.9%) of them were boys and CDE was right-sided in 73 (64.6%) cases. Eighty-one patients (72.3%) were symptomatic, with respiratory issues in 76 (67.9%) cases, gastro-intestinal symptoms in 15 (13.4%) cases, and orthopedic abnormalities in nine (8.8%) cases. Respiratory impairment consisted in obstructive symptoms in 32.6% of the cases (asthma, bronchiolitis, chronic coughing), in respiratory distress or dyspnea in 25.0% of the cases, in recurrent respiratory tract infections in 15.2% and 4 (5.1%) were discovered on imaging exams carried out for unrelated reasons. Among digestive symptoms, failure to thrive and dysphagia or vomiting were reported in 5 cases each, and GERD occurred in 4 cases. Orthopedic abnormalities consisted in chest wall deformity or asymmetry in 5 cases, and scoliosis in 4 cases; they were associated to respiratory symptoms in five patients. On chest radiographs, the diaphragmatic dome was at a median 6^th^ rib level [5–7]. Imaging was completed by CT scan for 64 (57.1%) patients and by MRI for 10 (8.9%) patients, showing an inferior pulmonary lobe atelectasis in 29 cases (39.2%), 2 or 3-lobe atelectasia in four cases (5.4%), cardiac deviation in three cases (4.1%), and an associated pulmonary malformation in two cases (2.7%).

### 2) Description of surgical techniques

The median delay before surgery was 35 [5–120] days. A diaphragmatic plication was realized in 105 out of 112 patients (93.8%), consisting of a central plication as described by Patrini *et al.* [9] in 82 (73.2%) cases and a flag plication [3] in 23 (20.5%) patients; seven patients (6.3%) had a resection of the central fibrous part of the cupola and a plasty of the diaphragmatic muscle. Non-absorbable sutures were used in 99 (88.4%) patients, absorbable sutures in seven (6.3%), staples and prosthesis in two (1.8%) patients each. Plication was performed abdominally in 28 (25.0%) patients, including 12 (10.7%) laparotomies and 16 (14.3%) laparoscopies, and using a thoracic approach in the remaining 84 patients, comprising 15 (13.4%) postero-lateral thoracotomies (PLT), 64 (57.1%) video-assisted thoracoscopies (VATS) and five (4.5%) robot-assisted thoracoscopies (RATS) (Fig. 1). There was no correlation between the surgical approach and the plication technique or the material used for the intervention (Suppl. data, tables 1 - 3).

**Fig. 1:**
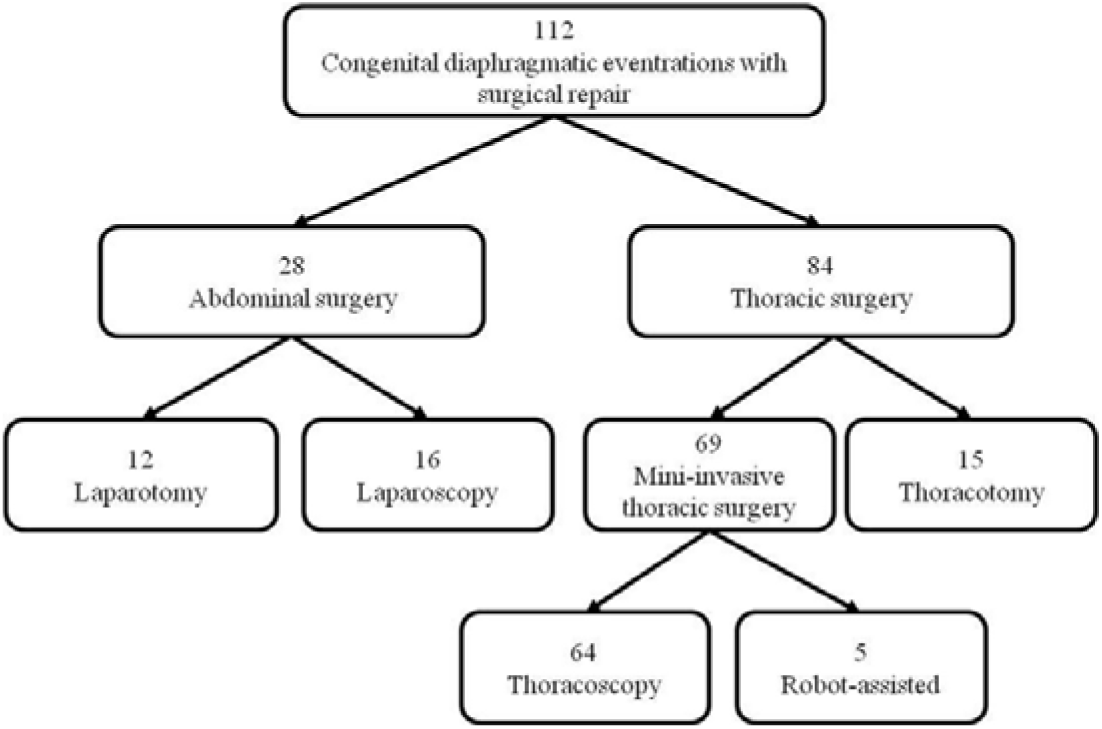
Flowchart of the cohort of patients operated on for congenital diaphragmatic eventration. Patients have been grouped according to the surgical approach used.

### 3) Comparative analysis of surgical approaches and outcomes

Analysis of preoperative data (Table 1) showed that the median age at diagnosis was not different between the three groups of patients divided according to surgical approach (abdominal approach, PLT, and MITS), nor the level of premature births. It also showed that there was no late diagnosis in the PLT group, in contrast to a rate of over 30% in the other groups. Patients operated on using PLT only presented right-sided CDE and none of them suffered from digestive or orthopedic problems. There was no difference between the three groups with regard to the proportion of symptomatic patients, but patients with digestive symptoms were three times more likely to be operated on using an abdominal approach than a thoracic approach. Furthermore, patients who had been treated using an abdominal approach had half as many respiratory symptoms as those who had undergone a thoracotomy. The level of the diaphragmatic dome and the side of the CDE did not appear to be a factor in the choice of the surgical approach, nor did the patient’s age at the time of surgery or the first or second intention surgery. On multivariate analysis, none of these factors was found to predispose patients to one specific surgical approach (Suppl. data, tables 4 - 6).

**Table 1:**
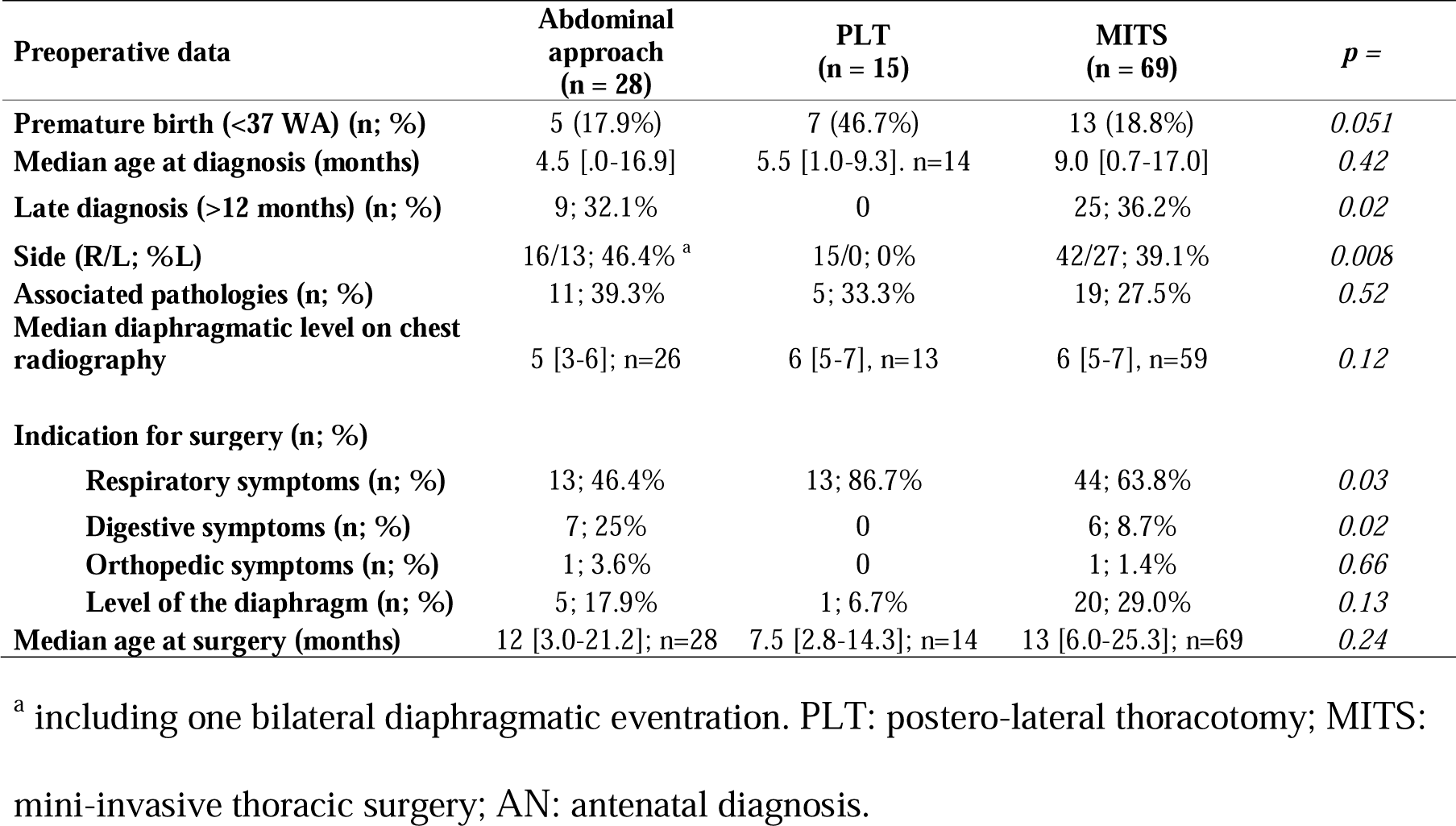
Comparison between surgical approaches for diaphragmatic plication.

Peri- and 30-day postoperative complication rates and types did not differ between groups (Table 2), but the duration of intravenous analgesia was three to four times longer in the PLT group (96 hours, versus 24 after abdominal approach and 36 after MITS, p = 0.0003), and its median 8-day hospital stay was longer than after MITS (4 days, p = 0.0007). The most frequent complications were infections in 10 cases out of 22 and pneumothorax in 7 cases; all of the latter occurred after MITS and were symptomatic and 4 out of them required exsufflation or iterative drainage.

**Table 2:** Comparison of peri and postoperative outcomes between surgical approaches.

| Perioperative data | Abdominal approach<br>(n = 28) | PLT<br>(n = 17)<sup>a</sup> | MITS<br>(n = 67)<sup>a</sup> | *p* = |
| --- | --- | --- | --- | --- |
| Perioperative complications (n; %) | 1; 3.6% | 0 | 8; 11.6% | 0.21 |
| Postoperative complications (n; %) | 2; 7.1% | 3; 17.6% | 17; 25.4% | 0.12 |
| Infections (n; %) | 1; 3.6% | 1; 5.9% | 8; 11.9% | 0.38 |
| Pneumothorax (n; %) | 0 | 0 | 7; 10.4% | 0.08 |
| Pleural effusion (n; %) | 1 (chylothorax) | 0 | 1; 1.5% | 0.57 |
| Respiratory distress / lobar atelectasis (n; %) | 0 | 1; 5.9% | 3; 4.5% | 0.48 |
| Occlusive syndrome (n; %) | 0 | 0 | 1; 1.5% | 0.34 |
| Secondary hiatal hernia (n; %) | 0 | 0 | 1; 1.5% | 0.34 |
| Subcutaneous emphysema (n; %) | 0 | 0 | 1; 1.5% | 0.34 |
| Claude-Bernard-Horner syndrom (n; %) | 0 | 0 | 1; 1.5% | 0.34 |
| Hemodynamic instability (n; %) | 0 | 0 | 2; 3.0% | 0.85 |
| Early diaphragmatic eventration or hernia recurrence (n; %) | 0 | 0 | 2; 3.0% | 0.85 |
| Not described (n; %) | 0 | 1; 5.9% | 0 | 0.94 |
| Persistent respiratory symptoms (n; %) | 4; 14.3% | 4; 23.5% | 7; 10.4% | 0.36 |
| Median use of IV analgesics (hours) | 24 [4.5-84.0] | 96 [72-138] | 36 [12-48] | 0.0003 |
| Median hospital stay (days) | 5 [2-9.5] | 8 [4.8-17] | 4 [3-5] | <0.0001 |

<sup>a</sup> Integration of the two cases of converted mini-invasive thoracic surgery (MITS) in the
postero-lateral thoracotomy (PLT) group for postoperative surgical results.

With a median follow-up of more than two years, there was no difference between groups concerning the persistence of symptoms or complication rates (Table 3). Seven deaths occurred, none of them directly related to CDE but rather to the underlying conditions and pathologies of the patients. Among the thoracic approach groups, nine patients (10.7%) underwent another surgery, consisting in an iterative plication for one patient in the PLT group and seven repeated plications or postoperatively-acquired diaphragmatic hernia cures in the MITS group. In the later, one patient had another surgery for hiatal hernia repair. No pre- or perioperative factor was predictive for the risk of recurrence on bi- and multivariate analysis (Suppl. data, table 7). On chest radiographs, the level of the cupola was improved by 3.5 costal height in the PLT group, higher than the two-point improvement on the other groups (p = 0.043), and there were no pulmonary or orthopedic abnormalities.

**Table 3:** Comparison of long-term outcomes between surgical approaches for diaphragmatic plication.

| Follow-up data | Abdominal approach<br>(n = 28) | PLT<br>(n = 17)<sup>a</sup> | MITS<br>(n = 67)<sup>a</sup> | *p* = |
| --- | --- | --- | --- | --- |
| Median follow-up (months) | 25 [7-62] | 37 [14-73] | 30 [11.5-56.5] | 0.55 |
| Symptoms (n; %) | 5; 17.9% | 5; 29.4% | 15; 22.4% | 0.66 |
| Respiratory (n; %) | 2; 7.1% | 4; 23.5% | 10; 14.9% | 0.30 |
| Digestive (n; %) | 0 | 2; 11.8% | 2; 3.0% | 0.46 |
| Orthopedic (n; %) | 3; 10.7% | 0 | 6; 9.0% | 0.40 |
| Overall complication rate (n; %) | 5; 17.9% | 4; 23.5% | 10; 14.9% | 0.69 |
| Secondary hospital stay (n; %) | 2; 7.1% | 2; 11.8% | 10; 1.5% | 0.58 |
| Iterative surgery (n; %) | 0 | 1; 5.9% | 8; 11.9% | 0.14 |
| Overall mortality (n; %) | 1; 3.6% | 3; 17.6% | 2; 3.0% | 0.05 |
| Chest radiography | 27; 96.4% | 15; 100% | 64; 95.5% | 0.44 |
| Median diaphragmatic level (back rib) | 8 [7-9] | 9.5 [8-10] | 9 [8-9] | 0.046 |
| Median difference in cupola level<br>(number of ribs) | 2 [1.5-3] | 3.5 [2-4] | 2 [1-3] | 0.043 |
| Improvement (n; %) | 10; 90.9% | 14; 100% | 45; 88.2% | 0.40 |
| Stability (n; %) | 1; 9.1% | 0 | 5; 9.8% | 0.48 |
| Aggravation (n; %) | 0 | 0 | 1; 2% | 0.78 |
| De novo pulmonary abnormalities (n; %) | 2; 7.4% | 0 | 3; 4.7% | 0.56 |
| De novo orthopedic abnormalities (n; %) | 0 | 0 | 5; 7.8% | 0.18 |

Long-term outcomes comparison between abdominal approach, postero-lateral thoracotomy
(PLT) and mini-invasive thoracic surgery (MITS). <sup>a</sup> Integration of the two cases of MITS in the PLT group for postoperative surgical results.

### 4) Robot-assisted thoracoscopic plication

Five patients, aged from three to120 months (median 29 [11.5 – 76.5]) at surgery, underwent RATS for CDE. Their preoperative characteristics did not differ from those of the 64 other patients operated on by VATS (Table 4). Indication to surgery was respiratory and/or digestive symptoms for three of them, an associated pulmonary sequestration for one, and an important elevation of the cupola (4^th^ rib) for the last one. Compared to VATS patients, diaphragmatic plication was the procedure that was performed the most (four patients) and resection-suturing of the diaphragm for the last case, but the perioperative use of absorbable suture was higher in the RATS group (40% versus 3%, p = 0.02), including the case of resection-suturing of the diaphragm. The risk of perioperative complication seemed identical, but the type of complications differed, with four pulmonary/pleural injuries and one tracheal tube dislodgement in the VATS group and two hepatic injuries with RATS, discovered during surgery by blood effusion, and conservatively treated without further clinical consequences. Median intravenous analgesia duration and hospital stay did not differ either. There was one recurrence of eventration after RATS and seven (11%, ns) after VATS, all of which required surgery to redo the procedure. After a median 48-month survey, symptoms remained stable or appeared in all five RATS patients, while they had disappeared for 35 (56.5%) VATS patients (p = 0.02). The radiographic level of the diaphragmatic cupola was improved in 40 (87%) patients following VATS and in all RATS patients, but orthopedic deformations had appeared in four (6%) and two (40%, p = 0.06) VATS and RATS patients, respectively.

**Table 4:** Pre- and perioperative comparison between video-assisted thoracoscopy and robot-assisted thoracoscopy.

|  | **VATS**<br>**(n=64)** | **RATS**<br>**(n=5)** | *p* = |
| --- | --- | --- | --- |
| **Preoperative data** |  |  |  |
| Mean term of birth (WA) | 37.9 +/- 2.9 | 38.8 +/- 1.5 | 0.84 |
| Median age at diagnosis (months) | 9.0 [1.0-17.0] | 4.0 [0.0-22.5] | 0.6 |
| Sex (%M) | 43; 67.2% | 3; 60% | 1 |
| Side of CDE (%G) | 24; 37.5% | 3; 60% | 0.37 |
| Symptoms | 46; 71.9% | 3; 60% | 0.62 |
| - respiratory | 45; 70.3% | 2; 40% | 0.32 |
| - digestive | 7; 10.9% | 2; 40% | 0.12 |
| - orthopedic | 8; 12.5% | 0 | 1 |
| Diaphragmatic level on chest radiograph (rib) | 6 [5-7] | 6 [4.5-7] | 0.51 |
| CT scan pulmonary abnormalities | 15; 23.4% | 1; 20% | 0.59 |
| **Perioperative data** |  |  |  |
| Median age at surgery (months) | 13.0 [6.0-23.3] | 29.0 [11.5-76.5] | 0.13 |
| Intervention: |  |  |  |
| - central plication | 49; 76.6% | 2; 40% | 0.11 |
| - flag plication | 13; 20.3% | 2; 40% | 0.30 |
| - excision-suturing | 2; 3.1% | 1; 20% | 0.20 |
| Suturing material: |  |  |  |
| - non absorbable | 58; 90.6% | 3; 60% | 0.10 |
| - absorbable | 2; 3.1% | 2; 40% | 0.02 |
| - staples | 2; 3.1% | 0 | 0.54 |
| - prosthesis | 1; 1.6% | 0 | 0.67 |
| Perioperative complications | 6; 9.4% | 2; 40% | 0.1 |
| Median length of IV analgesia (hours) | 24 [7.5-48] | 48 [39-66] | 0.17 |
| Median hospital stay (days) | 4 [3-5] | 5 [4.3-6.5] | 0.22 |
| **Outcomes** |  |  |  |
| Median age at the end of follow-up (months) | 48 [23-70] | 60 [29-91] | 0.85 |
| Median follow-up (months) | 28.8 [11.3-52.8] | 56.0 [10.5-65.0] | 0.59 |
| Surgical complications | 17; 27.4% | 0; 0% | 0.32 |
| Recurrence of eventration | 6; 9.7% | 1; 20% | 0.55 |
| Symptomatic patients | 11; 17.7% | 4; 80% | 0.01 |
| - Respiratory tract | 8; 12.9% | 2; 40% | 0.16 |
| - GERD | 1; 1.6% | 1; 20% | 0.14 |
| - Orthopedic | 4; 6.5% | 2; 40% | 0.06 |
| Diaphragmatic level on chest radiograph (rib) | 9 [8-9] | 9 [8.5-9] | 0.71 |
| Radiographic improvement | 40; 87% | 5; 100% | 1 |

VATS = video-assisted thoracoscopy; RATS = robot-assisted thoracoscopy; WA = weeks of
amenorrhea; CDE = congenital diaphragmatic eventration; IV = intra-venous; GERD =
gastro-esophageal reflux.

## Discussion

With 112 patients operated on for CDE during the past 12 years, our series is one of the most important to date and allows us to provide a reliable description of current trends in surgical care. The most widely used technique was the central plication, performed in 72% of the cases, followed by the “flag plication” technique in 20%. Excision-suturing of the cupola was only performed in few cases, maybe linked to the perioperative difficulty of accurately distinguishing between CDE with a congenital diaphragmatic hernia and a hernia sac. Furthermore, non-absorbable sutures were privileged to other closing means. On the whole, no difference in terms of efficiency or complications, including recurrence of diaphragmatic eventration, was found between the two plication techniques. Very few studies have addressed this topic. Bawazir *et al.*[10] compared interruptive sutures and the pleated technique in video-assisted thoracoscopy while Le Pimpec *et al.* [1] reported the flag plication as gold standard for thoracoscopy, with a similar conclusion to ours. No study has, however, specifically examined the usefulness of absorbable or permanent materials.

Video-assisted thoracoscopy was the most frequently used surgical approach, and is now challenged by the use of RATS, for which this study presents the first national cases of diaphragmatic plication. Thoracotomy and abdominal approaches tended to be used less, but if there was a trend to favor an abdominal approach in patients with digestive symptoms, no preoperative factor has been identified that guides the surgical approach and the latter was not related to the plication technique or the material used. In our series, MITS allowed for better and faster recovery than after PLT, slashing the length of hospital stays in half and decreasing the use of IV analgesics by three. Even if the quite long hospital stay after PLT may be partly related to a slightly higher amount of prematurity, these data agree with the findings previously published by Zhao *et al.*[6] where early postoperative recovery including mechanical ventilation, chest drainage time, and hospital stay was improved after thoracoscopy. After thoracoscopy, the lowering of the diaphragmatic cupola was certainly less important than after PLT, but the clinical resolution of symptoms during follow-up was the same in both groups.

There are very little data available on robot-assisted surgery for diaphragmatic eventration. A recent study by Bin Asaf *et al.* [11] including 18 adults patients proved its feasibility using thoracic and abdominal approaches, with a systematic improvement of pulmonary function test results. Studying children specifically, Slater *et al.*[12] in 2008 and Xu *et al.* [13] in 2020 have shown on two and nine patients, respectively, that robot-assisted diaphragmatic plication was feasible; just as in our five cases, they reported the same results pertaining to the efficacy of video-assisted surgery and without any additional complications. As Lampridis *et al.* [14], they report a shorter operative time due to greater ease during suturing, and an even better recovery. In our study, perioperative complications differed between RATS and VATS patients. There were two liver injuries with RATS, a known complication of diaphragmatic plication surgeries, which might be linked here to the absence of force feedback in robotic surgery, which causes the exact position of the hepatic dome to be underestimated. On the contrary, RATS and its 3D-improved visibility and enhanced ergonomy may help in preventing the pulmonary or pleural effusion encountered with VATS. It is worth noting that postoperative outcomes showed that more patients remained symptomatic after RATS than after VATS, with, in particular, two surgery-related chest wall abnormalities that might be linked to the larger diameter of robotic trocars.

Overall outcomes were satisfactory with 88.3% of immediate resolution of symptoms and 90% of diaphragmatic level improvement on chest radiographs. On the whole, most series describe prompt and durable improvement or resolution of symptoms after surgery in 91 to 100% of the cases [2,4,6,7,10], with 0 to 7% of incomplete repair or eventration recurrence [10,15,16], which is comparable to our 7.1% recurrence rate. Seven out of the eight recurrences of diaphragmatic eventration occurred following thoracoscopy. It may be compared to the higher recurrence rate already described after the thoracoscopic repair of congenital diaphragmatic hernia, even if all potential technical explanations do not fit the context of CDE [17,18]. Twenty-two patients (20%) experienced postoperative complications, mostly infections and pneumothorax that were medically treated, and only three were deemed to be serious (grades 3 – 4) according to the Clavien-Dindo classification [19]. Few publications specifically report on postoperative complications of diaphragmatic plication, which are close to the ones described after pulmonary surgery [3], associated with a higher risk of intra-abdominal organ injury [9]. A recent publication on RATS for diaphragmatic plication reported a similar 26.8% rate of postoperative complications [20].

The main weakness of our study is its retrospective and multicentric character, linked to the rarity of CDE, which does not allow for a prospective monocentric study design. The small size of the group does not permit us to entertain definite conclusions, particularly on the association analysis which were carried out for the purposes of gathering further information. Nevertheless, our study offers a good reflection of current surgical practices in treating CDE. It also shows technological adaptive changes in surgical approaches with the appearance of robotic surgery, which contributes to the options available to a pediatric surgeon seeking to offer the best treatment to their patients, thanks to technological innovations.

## Conclusion

Plication for congenital diaphragmatic eventration represents an efficient means to improve pulmonary and/or digestive symptoms and to normalize the level of the diaphragm on postoperative chest radiographs. Owing to a greater level of early postoperative recovery, mini-invasive techniques should be preferred over thoracotomy, but may expose patients to a somewhat higher risk of recurrence. An abdominal approach may be favored in cases of associated digestive surgery, on bilateral forms, or when the patient’s respiratory status does not allow for a thoracic approach. Robot-assisted thoracoscopy emerges as a valuable approach to diaphragmatic plication in children. Further studies are, however, required to determine whether or not it exposes patients to a higher rate of surgically induced chest wall deformations.

## Data Availability

All data produced in the present study are available upon reasonable request to the authors

## Declarations section

Dr Alzahrani, Dr Heng, Pr Khen-Dunlop, Dr Panait, Dr Hervieux, Dr Grynberg, Pr Abbo, Dr Hameury, Pr Lavrand, Dr Maillet, Dr Haffreingue, Dr Lehn, Dr De Napoli Cocci, Dr Habonimana, Pr Michel, Dr Montalva, Pr Ballouhey, Dr Fotso Kamdem, Dr Lecompte, Dr Liné, Dr Poupalou, Dr Meignan, Dr Deslandes, Pr Podevin and Dr Schmitt have no conflict of interest to disclose.

## Supplementary material

### PedDiaVen - Association analysis through logistic regression

#### 1) Postoperative outcomes associated with an abdominal or a thoracic surgical approach

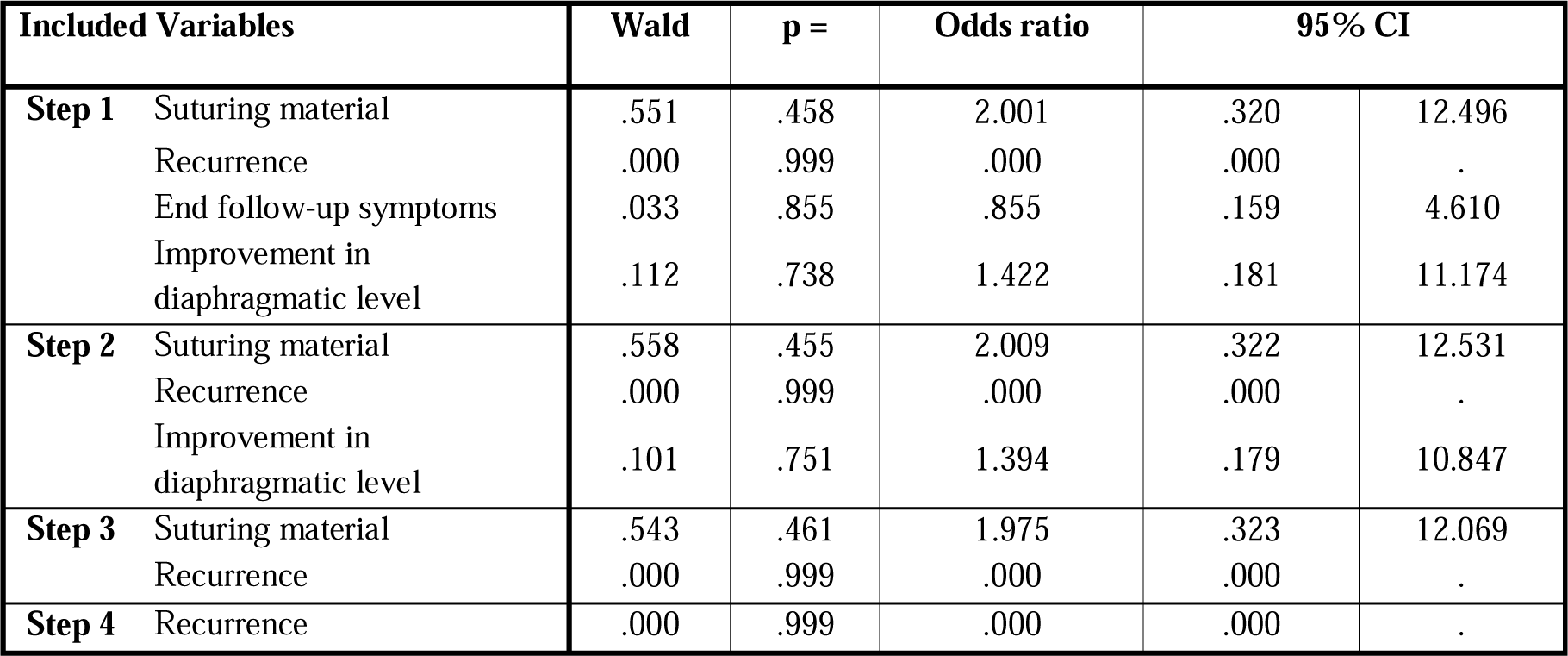

| Included Variables | Wald | p = | Odds ratio | 95% CI |  |
| --- | --- | --- | --- | --- | --- |
| **Step 1** Suturing material | .551 | .458 | 2.001 | .320 | 12.496 |
| Recurrence | .000 | .999 | .000 | .000 | . |
| End follow-up symptoms | .033 | .855 | .855 | .159 | 4.610 |
| Improvement in diaphragmatic level | .112 | .738 | 1.422 | .181 | 11.174 |
| **Step 2** Suturing material | .558 | .455 | 2.009 | .322 | 12.531 |
| Recurrence | .000 | .999 | .000 | .000 | . |
| Improvement in diaphragmatic level | .101 | .751 | 1.394 | .179 | 10.847 |
| **Step 3** Suturing material | .543 | .461 | 1.975 | .323 | 12.069 |
| Recurrence | .000 | .999 | .000 | .000 | . |
| **Step 4** Recurrence | .000 | .999 | .000 | .000 | . |

#### 2) Postoperative factors associated with postero-lateral thoracotomy (as compared to abdominal + mini-invasive thoracic surgery)

| Included Variables | Wald | p = | Odds ratio | 95% CI |  |
| --- | --- | --- | --- | --- | --- |
| **Step 1** Suturing material | .018 | .894 | 1.106 | .252 | 4.855 |
| Recurrence | .000 | .999 | 467175883.699 | .000 | . |
| End follow-up symptoms | .911 | .340 | .512 | .129 | 2.024 |
| Improvement in diaphragmatic level | 1.019 | .313 | 3.573 | .302 | 42.313 |
| **Step 2** Recurrence | .000 | .999 | 462390933.676 | .000 | . |
| End follow-up symptoms | .916 | .338 | .511 | .129 | 2.020 |
| Improvement in diaphragmatic level | 1.019 | .313 | 3.573 | .302 | 42.346 |
| **Step 3** Recurrence | .000 | .999 | 400698028.893 | .000 | . |
| Improvement in diaphragmatic level | .854 | .355 | 3.147 | .277 | 35.777 |
| **Step 4** Recurrence | .000 | .999 | 381839441.316 | .000 | . |

#### 3) Postoperative factors associated with mini-invasive thoracoscopic surgery (as compared to thoracotomy + abdominal approach)

| Included Variables |  | Wald | p = | Odds ratio | 95% CI |  |
| --- | --- | --- | --- | --- | --- | --- |
| **Step 1** | Suturing material | .183 | .669 | 1.327 | .363 | 4.858 |
|  | Recurrence | .000 | .999 | .000 | .000 | . |
|  | End follow-up symptoms | .417 | .518 | 1.484 | .448 | 4.912 |
|  | Improvement in diaphragmatic level | .384 | .536 | .578 | .102 | 3.274 |
| **Step 2** | Recurrence | .000 | .999 | .000 | .000 | . |
|  | End follow-up symptoms | .405 | .525 | 1.474 | .446 | 4.868 |
|  | Improvement in diaphragmatic level | .388 | .533 | .576 | .102 | 3.260 |
| **Step 3** | Recurrence | .000 | .999 | .000 | .000 | . |
|  | Improvement in diaphragmatic level | .317 | .574 | .611 | .110 | 3.395 |
| **Step 4** | Recurrence | .000 | .999 | .000 | .000 | . |

#### 4) Predisposing factors for an abdominal or a thoracic surgical approach

| Included Variables |  | Wald | p = | Odds ratio | 95% CI |  |
| --- | --- | --- | --- | --- | --- | --- |
| **Step 1** | Late diagnosis | .003 | .958 | 1.028 | .373 | 2.834 |
|  | Sex | .040 | .841 | 1.103 | .421 | 2.888 |
|  | Side | .615 | .433 | 1.479 | .556 | 3.937 |
|  | Associated abnormality | .491 | .483 | 1.410 | .539 | 3.689 |
|  | Respiratory symptoms | .955 | .329 | .624 | .243 | 1.606 |
|  | Digestive symptoms | .366 | .545 | 1.479 | .417 | 5.249 |
| **Step 2** | Sex | .045 | .832 | 1.108 | .429 | 2.861 |
|  | Side | .683 | .408 | 1.490 | .579 | 3.832 |
|  | Associated abnormality | .492 | .483 | 1.411 | .539 | 3.690 |
|  | Respiratory symptoms | .965 | .326 | .627 | .247 | 1.591 |
|  | Digestive symptoms | .379 | .538 | 1.485 | .422 | 5.222 |
| **Step 3** | Side | .660 | .416 | 1.477 | .576 | 3.785 |
|  | Associated abnormality | .513 | .474 | 1.420 | .544 | 3.706 |
|  | Respiratory symptoms | .955 | .328 | .629 | .248 | 1.594 |
|  | Digestive symptoms | .388 | .533 | 1.491 | .424 | 5.245 |
| **Step 4** | Side | .975 | .323 | 1.583 | .636 | 3.938 |
|  | Associated abnormality | .721 | .396 | 1.502 | .587 | 3.842 |
|  | Respiratory symptoms | .924 | .337 | .635 | .251 | 1.604 |
| **Step 5** | Side | .806 | .369 | 1.512 | .613 | 3.730 |
|  | Respiratory symptoms | 1.146 | .284 | .605 | .241 | 1.518 |
| **Step 6** | Respiratory symptoms | 1.505 | .220 | .567 | .229 | 1.404 |

#### 5) Preoperative factors associated with postero-lateral thoracotomy (as compared to abdominal + mini-invasive thoracic surgery)

| Included Variables |  | Wald | p = | Odds ratio | 95% CI |  |
| --- | --- | --- | --- | --- | --- | --- |
| **Step 1** | Late diagnosis | .000 | .998 | 345258340.173 | .000 | . |
|  | Sex | 1.117 | .291 | 2.200 | .510 | 9.494 |
|  | Side | 1.544 | .214 | 2.899 | .541 | 15.529 |
|  | Associated abnormality | .088 | .767 | 1.230 | .313 | 4.831 |
|  | Respiratory symptoms | 1.128 | .288 | .458 | .108 | 1.937 |
|  | Digestive symptoms | .054 | .817 | 1.315 | .129 | 13.400 |
|  | Neonatal surgery | .182 | .669 | 1.462 | .256 | 8.368 |
| **Step 2** | Late diagnosis | .000 | .998 | 347414180.063 | .000 | . |
|  | Sex | 1.167 | .280 | 2.232 | .520 | 9.578 |
|  | Side | 1.582 | .208 | 2.933 | .549 | 15.678 |
|  | Associated abnormality | .127 | .722 | 1.275 | .335 | 4.855 |
|  | Respiratory symptoms | 1.146 | .284 | .455 | .108 | 1.924 |
|  | Neonatal surgery | .158 | .691 | 1.421 | .252 | 8.012 |
| **Step 3** | Late diagnosis | .000 | .998 | 347848774.883 | .000 | . |
|  | Sex | 1.139 | .286 | 2.202 | .517 | 9.386 |
|  | Side | 1.497 | .221 | 2.810 | .537 | 14.702 |
|  | Respiratory symptoms | 1.279 | .258 | .439 | .105 | 1.829 |
|  | Neonatal surgery | .203 | .652 | 1.483 | .267 | 8.237 |
| **Step 4** | Late diagnosis | .000 | .998 | 324570840.350 | .000 | . |
|  | Sex | 1.024 | .312 | 2.098 | .500 | 8.806 |
|  | Side | 1.643 | .200 | 2.932 | .566 | 15.196 |
|  | Respiratory symptoms | 1.174 | .279 | .457 | .111 | 1.883 |
| **Step 5** | Late diagnosis | .000 | .998 | 355411695.750 | .000 | . |
|  | Side | 1.250 | .263 | 2.533 | .497 | 12.921 |
|  | Respiratory symptoms | 1.029 | .310 | .482 | .117 | 1.975 |
| **Step 6** | Late diagnosis | .000 | .998 | 324692018.869 | .000 | . |
|  | Side | 1.942 | .163 | 3.088 | .632 | 15.075 |
| **Step 7** | Late diagnosis | .000 | .998 | 410713908.205 | .000 | . |

#### 6) Preoperative factors associated with mini-invasive thoracoscopic surgery (as compared to thoracotomy + abdominal approach)

| Included Variables |  | Wald | p = | Odds ratio | 95% CI |  |
| --- | --- | --- | --- | --- | --- | --- |
| **Step 1** | Late diagnosis | 2.342 | .126 | .468 | .177 | 1.238 |
|  | Sex | .068 | .794 | .889 | .367 | 2.153 |
|  | Side | .048 | .827 | .904 | .364 | 2.245 |
|  | Associated abnormality | .261 | .610 | 1.260 | .519 | 3.060 |
|  | Respiratory symptoms | .010 | .919 | .955 | .396 | 2.303 |
|  | Digestive symptoms | .355 | .551 | 1.459 | .421 | 5.057 |
|  | Neonatal surgery | .195 | .659 | 1.335 | .370 | 4.817 |
| **Step 2** | Late diagnosis | 2.467 | .116 | .464 | .178 | 1.210 |
|  | Sex | .069 | .793 | .888 | .367 | 2.152 |
|  | Side | .040 | .841 | .913 | .376 | 2.218 |
|  | Associated abnormality | .282 | .595 | 1.269 | .527 | 3.052 |
|  | Digestive symptoms | .353 | .553 | 1.457 | .421 | 5.044 |
|  | Neonatal surgery | .187 | .665 | 1.325 | .371 | 4.738 |
| **Step 3** | Late diagnosis | 2.778 | .096 | .453 | .179 | 1.149 |
|  | Sex | .057 | .811 | .898 | .374 | 2.161 |
|  | Associated abnormality | .313 | .576 | 1.282 | .537 | 3.063 |
|  | Digestive symptoms | .319 | .572 | 1.417 | .423 | 4.744 |
|  | Neonatal surgery | .170 | .680 | 1.305 | .369 | 4.618 |
| **Step 4** | Late diagnosis | 2.881 | .090 | .448 | .177 | 1.132 |
|  | Associated abnormality | .304 | .581 | 1.277 | .535 | 3.047 |
|  | Digestive symptoms | .320 | .572 | 1.417 | .423 | 4.747 |
|  | Neonatal surgery | .207 | .649 | 1.336 | .383 | 4.660 |
| **Step 5** | Late diagnosis | 3.342 | .068 | .429 | .173 | 1.063 |
|  | Associated abnormality | .330 | .566 | 1.290 | .542 | 3.071 |
|  | Digestive symptoms | .266 | .606 | 1.372 | .412 | 4.566 |
| **Step 6** | Late diagnosis | 3.151 | .076 | .446 | .183 | 1.088 |
|  | Associated abnormality | .457 | .499 | 1.342 | .572 | 3.150 |
| **Step 7** | Late diagnosis | 3.146 | .076 | .447 | .184 | 1.088 |

#### 7) Risk factors for congenital diaphragmatic eventration recurrence

| Included Variables | Wald | p = | Odds ratio | 95% CI |  |
| --- | --- | --- | --- | --- | --- |
| Late diagnosis | .209 | .648 | .509 | .028 | 9.213 |
| Sex | .000 | .998 | 309291785.373 | .000 | . |
| Side | .497 | .481 | 2.630 | .179 | 38.660 |
| Associated abnormality | .209 | .647 | 2.136 | .083 | 55.040 |
| Type of surgical approach | 2.381 | .123 | 7.648 | .577 | 101.368 |
| Neonatal surgery | .000 | .999 | 60292654.945 | .000 | . |
| Suturing material | .724 | .395 | 2.002 | .405 | 9.909 |
| End follow-up symptoms | .137 | .711 | .610 | .045 | 8.358 |
| Improvement in diaphragmatic level | .243 | .622 | 2.796 | .047 | 166.119 |

## Notes

**Financial support disclosure:** This research received no specific grants from funding agencies in the public, commercial, or not-for-profit sectors.

### Competing Interest Statement

The authors have declared no competing interest.

### Funding Statement

This study did not receive any funding

### Author Declarations

The ethics committee of the University Hospital Center of Angers gave ethial approval for this work

### Summary of Updates

Some descriptions have been provided in the material and methods section; tables have been simplified.

